# Predicting response and resistance to immune checkpoint blockade and surgery in melanoma patients

**DOI:** 10.1101/2023.10.05.23296626

**Authors:** Guillaume Mestrallet

## Abstract

Melanoma remains a formidable clinical challenge, claiming the lives of 60,000 patients annually. Current therapeutic modalities encompass surgical intervention and immune checkpoint inhibitors (ICB). Nevertheless, the efficacy of ICB varies, necessitating the need to anticipate response and resistance outcomes, while also considering alternative approaches, such as surgical interventions coupled with autologous skin grafts. In pursuit of these objectives, we conducted a comprehensive analysis involving seven melanoma patient cohorts subjected to four distinct ICB treatments. Remarkably, our findings revealed varying response rates: 29% for Nivolumab, 43% for Pembrolizumab, 20% for Ipilimumab, and an encouraging 62.5% for the combination of Pembrolizumab and Ipilimumab. This underscores the superior clinical outcomes associated with anti-PD1+anti-CTLA4 therapy. Intriguingly, responders to Pembrolizumab and Ipilimumab exhibited distinct immunological characteristics, characterized by an augmentation in Th1 and M1 macrophages, alongside a reduction in CD4+ T cell infiltration. This phenomenon coincided with the upregulation of antigen presentation genes (HLA, CD80), heightened pro-inflammatory cytokine production (CCL5, CXCL9, CXCL10), and enhanced T cell responses. Furthermore, based on these response profiles, we have developed predictive software to forecast individual patient responses to available checkpoint inhibitor combinations. This innovative tool also facilitates precise calculations for the extent of melanoma resection required during surgery, graft sizing, and the determination of the necessary autologous skin cell resources. In conclusion, our approach advocates for tailored therapies, leveraging patient-specific attributes and computational predictions to enhance clinical outcomes following immunotherapy and surgical interventions. This strategy holds promise for advancing melanoma treatment paradigms.

## Introduction

Approximately 60,000 patients succumb to melanoma annually (1). Existing treatment modalities encompass immune checkpoint inhibitors (ICB), chemotherapy, or surgery (2). Clinical trials have explored various immune checkpoints, including anti-PD1 agents (Pembrolizumab or Nivolumab), anti-CTLA4 (Ipilimumab), and combinations of anti-CTLA4 with anti-PD-1 (Nivolumab or Pembrolizumab and Ipilimumab) (3–9). Immune responses under steady-state conditions or following ICB administration hinge upon antigen presentation to T cells, mediated by human leukocyte antigens (HLA) and related cell-surface molecules (MICA, MICB, and costimulation molecules) (10). T cell responses also entail the expression of multiple genes promoting T cell proliferation, cytotoxicity, and maintenance, such as BTN3A, TNFRSF, PRF1, and CD27 (11–14).

Crucially, not all patients respond to ICB, and immune resistance remains a challenge (3–9). Immune resistance may be attributed to the expression of diverse immune checkpoints and immunosuppressive pathways in melanoma patients, including PD-L1, TIM3, PD1, CTLA4, LAG3, PDL2, TIGIT, and HLA-G (2,10).

Therefore, it is imperative to conduct a meta-analysis of melanoma patient cohorts to elucidate the mechanisms underpinning immune response and resistance to different therapies, aiming to enhance the prediction of responses to ICB and surgery. Meta-analysis of data from multiple cohorts may facilitate the identification of optimal targets or combination therapies to improve patient outcomes and overcome resistance mechanisms. The development of software will automate and enhance the precision of response prediction to ICB and surgery, thus improving diagnosis and subsequent therapeutic strategies, including the preparation of autologous skin grafts following surgery (15).

## Material and methods

### RNAseq datasets and selection of cohorts

Patient cohort was selected using the CRI iAtlas Portal (16). We selected the following RNAseq datasets for melanoma patients : Gide 2019 - SKCM, PD-1 +/- CTLA4 (7), Hugo 2016 - SKCM, PD-1 (9), Liu 2019 - SKCM, PD-1 (5), Riaz 2017 - SKCM, PD-1 (8), Van Allen 2015 - SKCM, CTLA-4 (6), Chen 2016 - SKCM, Anti-CTLA4 (3), Prat 2017 - HNSC, LUAD, LUSC, SKCM, Anti-PD-1 (4). We used the following group filters : Responder, Drug and SKCM. Responders are defined as patients with mRECIST of Partial Response or Complete Response, whereas Non-Responders are those with Progressive Disease or Stable Disease. Then, we used the ICI Analysis Modules. The current version of the iAtlas Portal was built in R using code hosted at https://github.com/CRI-iAtlas/iatlas-app. Assayed samples were collected prior to immunotherapy.

### Clinical description of patients

Merged datasets according to drug therapy are described in **Table 1**. For patients that received Ipilimumab, targeting CTLA4, there are 33 (80%) non responders and 8 (20%) responders, 2 (5%) with complete response, 6 (15%) with partial response, 26 (63%) with progressive disease and 7 (17%) with stable disease. For patients that received Ipilimumab and Nivolumab, targeting PD1 and CTLA4, there are 3 (33%) non responders and 6 (67%) responders, 0 with complete response, 6 (67%) with partial response, 3 (33%) with progressive disease and 0 with stable disease. For patients that received Ipilimumab and Pembrolizumab, targeting PD1 and CTLA4, there are 12 (37.5%) non responders and 20 (62.5%) responders, 13 (40,625%) with complete response, 7 (21.875%) with partial response, 6 (18.75%) with progressive disease and 6 (18.75%) with stable disease. For patients that received Nivolumab, targeting PD1, there are 114 (71%) non responders and 46 (29%) responders, 15 (9.375%) with complete response, 31 (19.375%) with partial response, 75 (46.875%) with progressive disease and 39 (24.375%) with stable disease. For patients that received Pembrolizumab, targeting PD1, there are 78 (57%) non responders and 59 (43%) responders, 15 (11%) with complete response, 44 (32%) with partial response, 58 (42%) with progressive disease and 20 (15%) with stable disease.

**Table 1).** Response of patients with melanoma to immune checkpoint blockade.

| Drug | Ipilimumab | Ipilimumab and Nivolumab | Ipilimumab and Pembrolizumab | Nivolumab | Pembrolizumab |
| --- | --- | --- | --- | --- | --- |
| Target | CTLA4 | CTLA4 and PD1 | CTLA4 and PD1 | PD1 | PD1 |
| Non responders (%) | 80 | 33 | 37,5 | 71 | 57 |
| Responders (%) | 20 | 67 | 62,5 | 29 | 43 |
| Complete response (%) | 5 | 0 | 40,625 | 9,375 | 11 |
| Partial response (%) | 15 | 67 | 21,875 | 19,375 | 32 |
| Progressive disease (%) | 63 | 33 | 18,75 | 46,875 | 42 |
| Stable (%) | 17 | 0 | 18,75 | 24,375 | 15 |

### Immune landscape of cancer in iAtlas

The initial release of iAtlas provided a resource to complement analysis results from The Cancer Genome Atlas (TCGA) Research Network on the TCGA data set comprising over 10,000 tumor samples and 33 tumor types (The Immune Landscape of Cancer; here referred to as Immune Landscape) (17).

### Statistics

Statistical significance of the observed differences was determined using the independent Wilcoxon t-Test with multiple sample correction. All data are presented as mean±SEM. The difference was considered as significant when the p value was below 0.05. * : p<0.05.

### Software development to predict response to therapy

The probability to respond to each immune checkpoint combination therapy is calculated as indicated in **Table 1**. Patients from 7 cohorts received anti-CTLA4 and/or anti-PD-1 following melanoma. After pooling the cohorts, response and recist response were calculated according to the drug used for therapy. n=41 for Ipilimumab, n=9 for Ipilimumab and Nivolumab, n=32 for Ipilimumab and Pembrolizumab, n=160 for Nivolumab and n=137 for Pembrolizumab.

We used the same parameters to calculate the size of the skin surface to remove by surgery and the size of the graft to produce, the time and the cells necessary to produce the graft according to patient characteristics as explained in our previous work on severe burn management and agronomy (15,18). Briefly, the software, coded using python, html, css, mysql and django, allows the registered clinician to diagnose a new patient or access the diagnosis of a registered patient by indicating their medical identifier in a form. The software calculates the size of the melanoma on the head and neck, the lower limbs, the upper limbs, the external genitalia, the posterior surface of the trunk and the anterior surface of the trunk. Importantly, the percentage of the total area of the body that these different areas represent varies depending on the age, the sex, the size and the weight of the patient. Informing the patient’s characteristics therefore allows these different areas to be assigned an appropriate value. Each value represents a percentage of the total body surface based on the characteristics of the patient. Then the calculation of the total area of the patient’s body from his weight and height is done using the Du Bois formula (19).

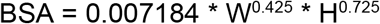

With BSA = Body Surface Area in m^2^, W = weight in kg and H = height in cm.

After form completion, the software calculates the following variables (**Table 2**) and displays the patient diagnostic.

**Table 2).** Variables calculated by the software for diagnosis and treatment.

|  |
| --- |
| Probability to respond to immunotherapy (%) |
| Size of the patient melanoma (% of total body area) |
| Graft surface to prepare (in cm<sup>2</sup>) |
| Surface of the biopsy to be taken to prepare the autologous graft (in cm<sup>2</sup>) |
| Duration of amplification and cell culture to obtain the graft (in days) |
| Graft preparation protocol |

The percentage of the total surface of the body to which each zone corresponds is calculated according to age, with the rule of Wallace or that of Lund and Browder (20–22). The size of the graft to be prepared is calculated by summing the melanoma areas. The size of the skin biopsy to be taken and the graft preparation protocol is determined by previous clinical observations (23). The choice was made to fix the maximum size of the sample at 4 cm^2^ to avoid unduly altering the unaffected areas of the patient. Keratinocyte amplification and graft production is performed as previously described (10,15,24,25). The number of cells needed to prepare the graft and the size of the biopsy to be taken to obtain them depends on the preparation time of the graft chosen. We used two formulas previously described to calculate them (15).

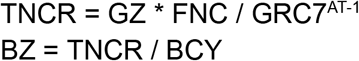

With TNCR = total number of cells required for the first amplification during the first 7 days, BZ = biopsy size (cm^2^), GZ = graft size (cm^2^), FNC = Final number of cells to seed per cm^2^ 7 days before grafting. At this stage, 7 days before the transplant, it goes from cell amplification to graft production. GRC7 = growth rate of cells in 7 days (average number of cells obtained from a single cell), AT = amplification time (weeks), BCY = biopsy cell yield (number of cells extracted from a biopsy cm^2^). Here FNC is equal to 10,000 cells/cm^2^, GRC7 of keratinocytes is equal to 17 cells and BCY is equal to 1,250,000 cells/cm^2^.

## Results

### Response and overall survival of melanoma patients according to immune checkpoint blockade in multiple cohorts

We examined the response and overall survival of melanoma patients who underwent immune checkpoint therapy targeting CTLA4 and/or PD1 in seven cohorts. We aggregated the results based on the response status to each checkpoint combination.

For patients who received Ipilimumab, targeting CTLA4, there were 33 (80%) non-responders and 8 (20%) responders, including 2 (5%) with a complete response, 6 (15%) with a partial response, 26 (63%) with progressive disease, and 7 (17%) with stable disease (**Table 1**). In the group of patients treated with Nivolumab, targeting PD1, there were 114 (71%) non-responders and 46 (29%) responders, with 15 (9.375%) having a complete response, 31 (19.375%) a partial response, 75 (46.875%) experiencing progressive disease, and 39 (24.375%) showing stable disease. Among patients treated with Pembrolizumab, targeting PD1, there were 78 (57%) non-responders and 59 (43%) responders, with 15 (11%) achieving a complete response, 44 (32%) a partial response, 58 (42%) experiencing progressive disease, and 20 (15%) showing stable disease.

In the case of patients who received Ipilimumab and Nivolumab, targeting PD1 and CTLA4, there were 3 (33%) non-responders and 6 (67%) responders, with no complete responses, 6 (67%) showing a partial response, and 3 (33%) experiencing progressive disease. No cases of stable disease were observed. Among patients who received Ipilimumab and Pembrolizumab, targeting PD1 and CTLA4, there were 12 (37.5%) non-responders and 20 (62.5%) responders, with 13 (40.625%) achieving a complete response, 7 (21.875%) with a partial response, 6 (18.75%) showing progressive disease, and 6 (18.75%) experiencing stable disease.

For patients receiving monotherapy targeting PD1 or CTLA4 without response, overall survival remained below 20% (**Figure 1**). Conversely, patients responding to monotherapy exhibited an overall survival rate between 50% and 90%. Patients receiving combination therapy targeting PD1 and CTLA4 without response displayed an overall survival rate around 50%. In contrast, patients responding to combination therapy demonstrated an overall survival rate around 100%. Overall, the most favorable clinical outcomes were observed among melanoma patients who received anti-PD1+anti-CTLA4 therapy. However, 30% to 40% of patients exhibited resistance to this immune checkpoint combination therapy.

**Figure 1).**
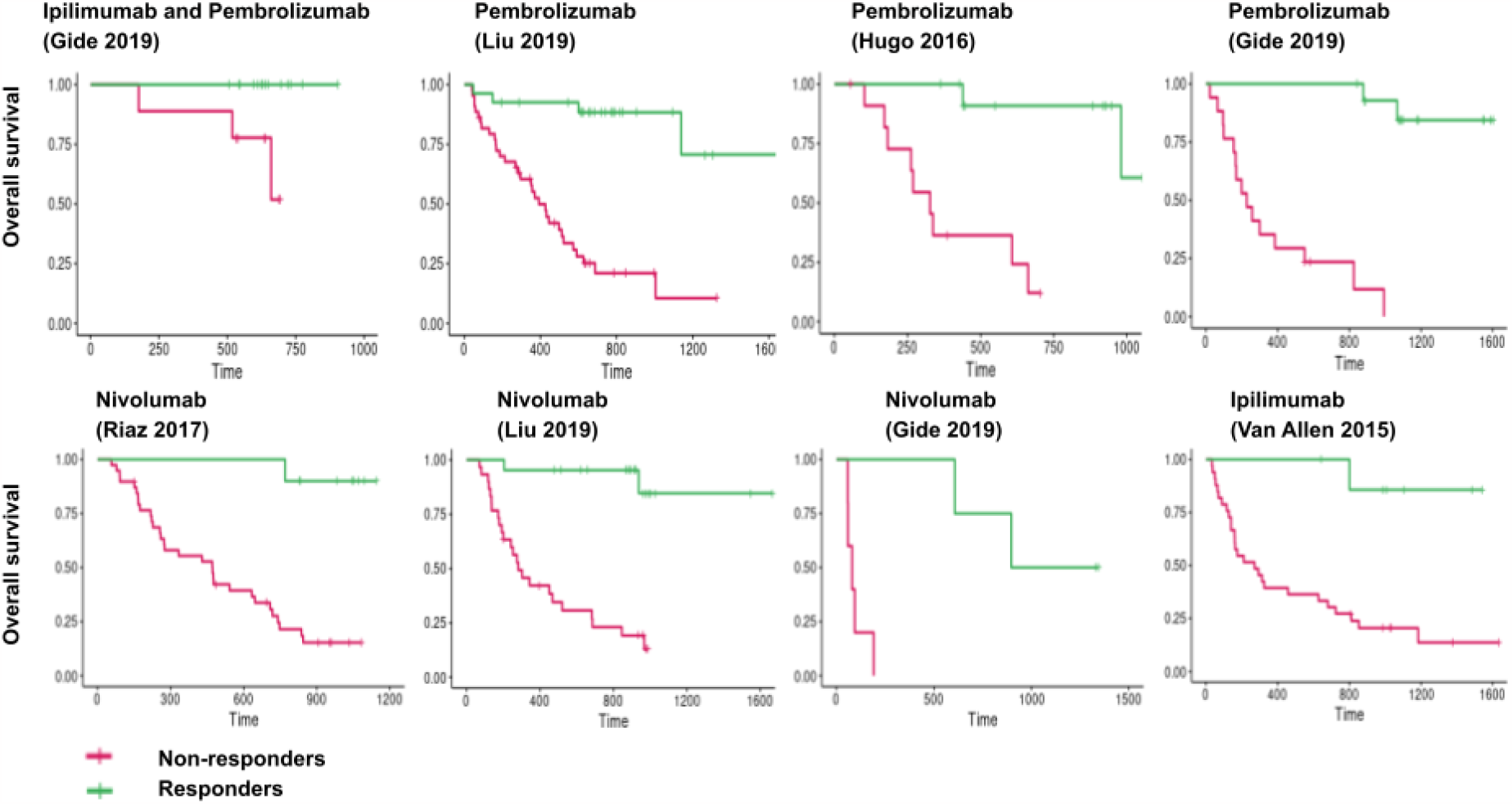
Overall survival of melanoma patients according to immune checkpoint blockade in multiple cohorts. Patients from 5 cohorts received anti-CTLA4 and/or anti-PD-1 following melanoma. Overall survival was calculated in these cohorts. n=41 for Ipilimumab, n=32 for Ipilimumab and Pembrolizumab, n=160 for Nivolumab and n=137 for Pembrolizumab.

### Immune response and resistance in melanoma patients following immune checkpoint blockade

We investigated the immune features associated with response and resistance to immune checkpoint therapy in melanoma patients. Analyzing immune infiltration using the CRI iAtlas in patients who received anti-PD1 and anti-CTLA4 (Ipilimumab and Pembrolizumab), we observed that non-responders had more CD4+ T cells, while responders had more Th1 and M1 macrophages (**Figure 2**). No significant differences were observed for other immune subsets.

**Figure 2).**
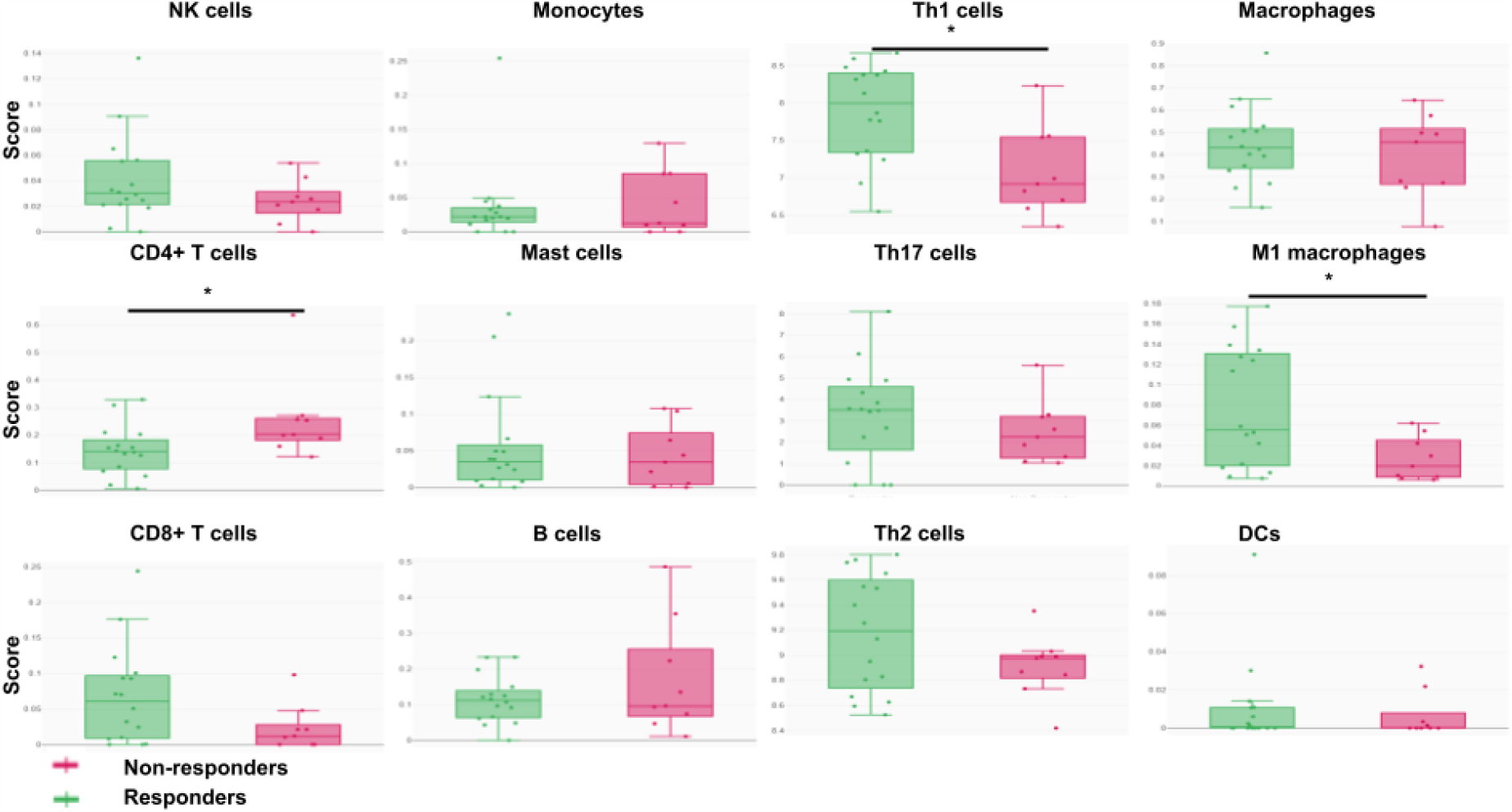
Immune infiltration in melanoma patients according to response to anti-PD1 + anti-CTLA4. 32 patients received anti-CTLA4 and anti-PD-1 following melanoma (Ipilimumab and Pembrolizumab). Immune infiltration is measured using CRI iAtlas. p<0.05, Wilcoxon t-test.

When examining the expression of immunomodulatory molecules, we found that responders expressed more MHC and costimulatory molecules (HLA-DRA, HLA-A, HLA-B, HLA-DRB1, HLA-DQA2, HLA-DQA1, HLA-DPA1, and CD80) compared to non-responders. This suggests that antigen presentation plays a crucial role in the anti-tumor immune response (**Figure 3**). Responders also exhibited higher expression of PD1 and TIGIT, indicating a potential increase in T cell exhaustion. We observed upregulation of genes related to T cell activation (BTN3A2), adhesion (ITGB2, ICAM1), cytotoxicity (PRF1), regulation (SLAMF7, IDO1), and maintenance (CD27) in responders. Additionally, responders showed increased expression of PD1 ligands PDL1 and PDL2, TNF signaling genes (TNFSF4, TNFSF9), and cytokines attracting T cells and other immune cells (CCL5, CXCL9, CXCL10). No differences were observed in the expression of other immunoregulatory molecules and immune checkpoints (including TIM3, LAG3, EDNRB, TLR4, ARG1, VSIR, CD40, TNFRSF, CD28, ICOS, VTCN1, CD70, CX3CL1, ENTPD1, GXMA, HMGB1, ICOSLG, VEGF, KIR, IFN/TGFB genes, interleukins, MICA, and other HLA genes).

**Figure 3).**
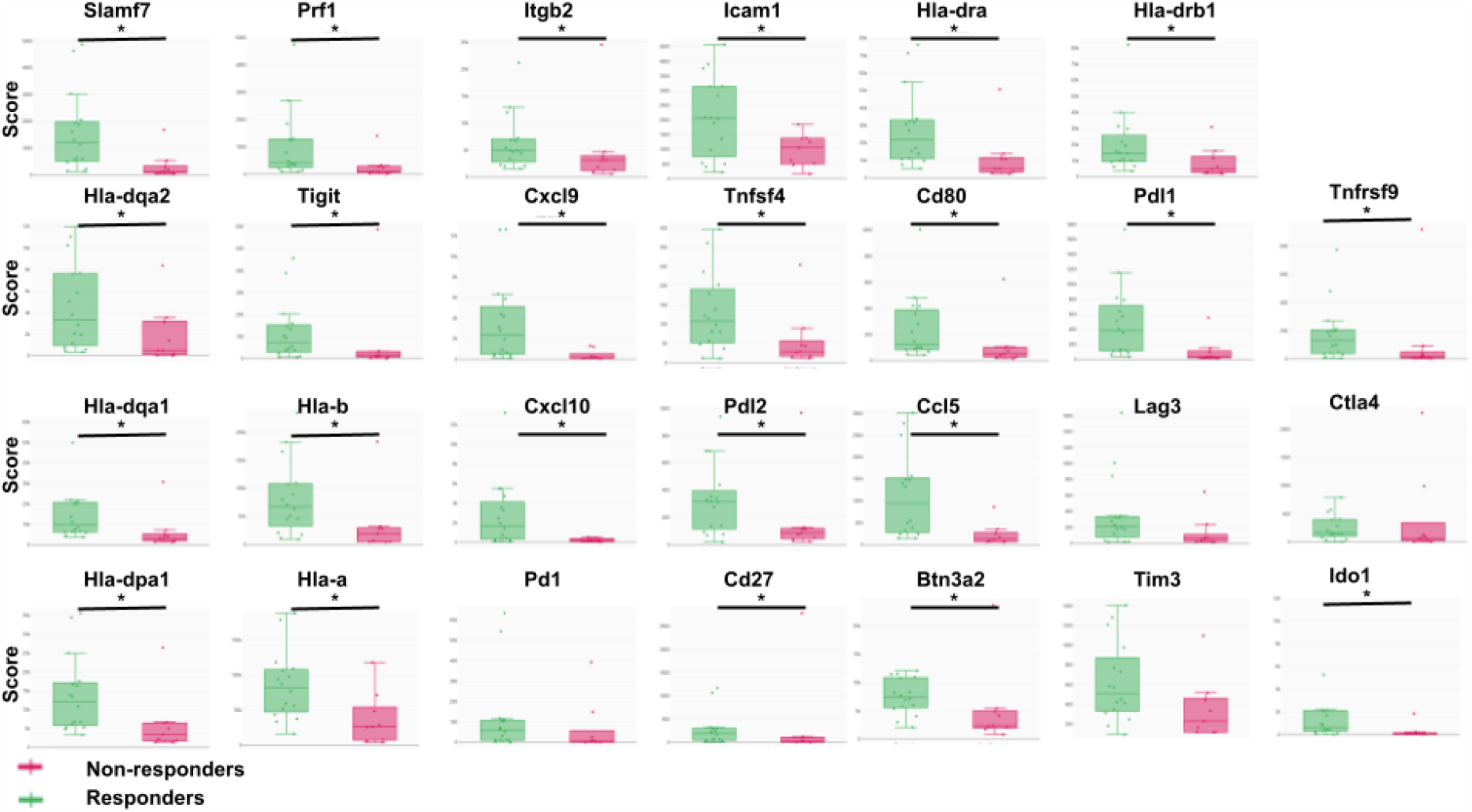
Immunomodulatory molecule expression in melanoma patients according to response to anti-PD1 + anti-CTLA4. 32 patients received anti-CTLA4 and anti-PD-1 following melanoma (Ipilimumab and Pembrolizumab). Immunomodulatory molecule expression is measured using CRI iAtlas. p<0.05, Wilcoxon t-test.

Interestingly, following Pembrolizumab monotherapy, differential immune infiltration was observed by B cells, memory B cells, and M1 macrophages, along with differential expression of IDO1, PDL1, HLA-DRB1, TIGIT, CTLA4, LAG3, and PD1 between responders and non-responders in the Gide cohort. However, such differences were not observed in the other cohorts (**Supplementary Figures 1/2**). Similarly, after Nivolumab monotherapy, differential expression of IDO1, CTLA4, and CD28 in the Riaz cohort, as well as CD40, HLA-A, and ICAM1 in the Liu cohort between responders and non-responders were noted, but again, not in the other cohorts. In the case of Ipilimumab monotherapy, we observed a higher presence of monocytes in responders. Overall, melanoma responders to checkpoint combination therapy were characterized by improved antigen presentation, macrophage, and T cell responses.

### Software prediction of patient response to immune checkpoint blockade and surgery

In accordance with the methods outlined in the study (**Figure 4**), we conducted predictions regarding the response of a hypothetical patient to immune checkpoint blockade and surgery. The calculated probability of the patient responding to Ipilimumab and Pembrolizumab is 62.5%, with a probability of achieving a complete response at 40.625%. The graft, required for the surgical procedure, can be generated within approximately 14 days from a 4 cm^2^ biopsy. The size of the graft to be produced corresponds to the dimensions of the melanoma to be removed during surgery, which, in this instance, is 2,067 cm^2^. The graft production protocol is provided, along with the requisite number of cells for each step.

**Figure 4).**
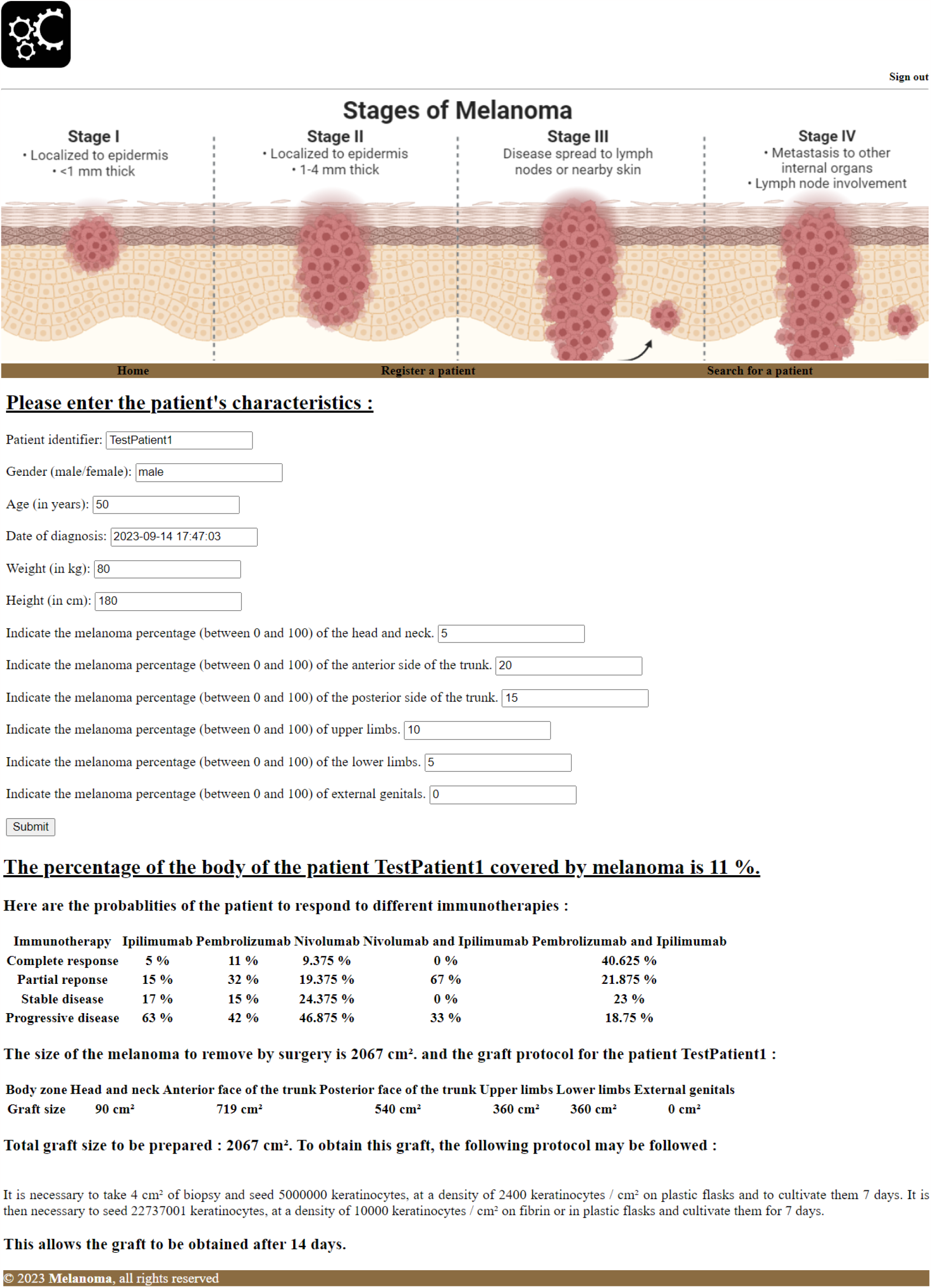
Software prediction of patient response to immune checkpoint blockade and surgery. Melanoma patient data collection form. Probability of the patient to respond to immune checkpoint blockade and size of the surgery to perform. Personalized graft production protocol for the patient following surgery.

## Discussion

By amalgamating data from seven cohorts, we observed that 20% to 67% of patients with melanoma responded to various ICB therapies. In more detail, 29% responded to the anti-PD1 agent Nivolumab, 43% responded to anti-PD1 Pembrolizumab, 20% responded to anti-CTLA4 Ipilimumab, and 62.5% responded to the combination of anti-CTLA4 with anti-PD-1 (Pembrolizumab and Ipilimumab). The most favorable clinical outcomes were evident in melanoma patients who received anti-PD1+anti-CTLA4 therapy.

Responders to Pembrolizumab and Ipilimumab exhibited distinctive characteristics, including an increase in Th1 cells and Macrophages M1, as well as a decrease in CD4+ T cell infiltration. These responders also demonstrated elevated antigen presentation (HLA, CD80) and heightened T cell responses (BTN3A2, ITGB2, ICAM1, PRF1, CD27). Furthermore, responders expressed a greater abundance of cytokines that attract T cells and other immune cells (CCL5, CXCL9, CXCL10). CXCL9 stimulates cytokine production by T cells and fosters the proliferation of Th1 cells (26). CXCL10, an inflammatory chemokine, facilitates immune responses by activating and recruiting T cells, eosinophils, monocytes, and NK cells (27). CCL5 plays a pivotal role in recruiting various leukocytes to inflammatory sites, including T cells, macrophages, eosinophils, and basophils (28).

However, no immune features associated with immune resistance were identified within these cohorts, and 37.5% of the patients continued to exhibit resistance even to combination therapy. Furthermore, no consistent immune features were shared across all cohorts in relation to response or resistance to monotherapies. Resistance mechanisms may be more influenced by cancer cells, as indicated by single-cell studies, and may be targeted by CDK4/6 inhibitors (29). Consequently, the stimulation of pathways identified in responders to multiple checkpoint therapies, such as antigen presentation and cytokine production, could potentially enhance the anti-tumor response mediated by Th1 cells and macrophages in non-responders.

Finally, we developed software to predict patient responses to immune checkpoint blockade and surgery, as detailed in the methods section. This software computes the probability of responding to each available checkpoint combination and facilitates improved calculations for the extent of tissue to be removed during surgery. Additionally, it estimates the number of days required to produce a graft from a 4 cm^2^ biopsy. This software provides a graft production protocol, specifying the number of cells required for each step, akin to the software used in the management of severe burns (15).

## Data Availability

Data available on cri iAtlas

## Acknowledgements

The author thanks Dr A. Pierga for the logo of the software.

## Supplementary figures

**Supplementary figure 1).**
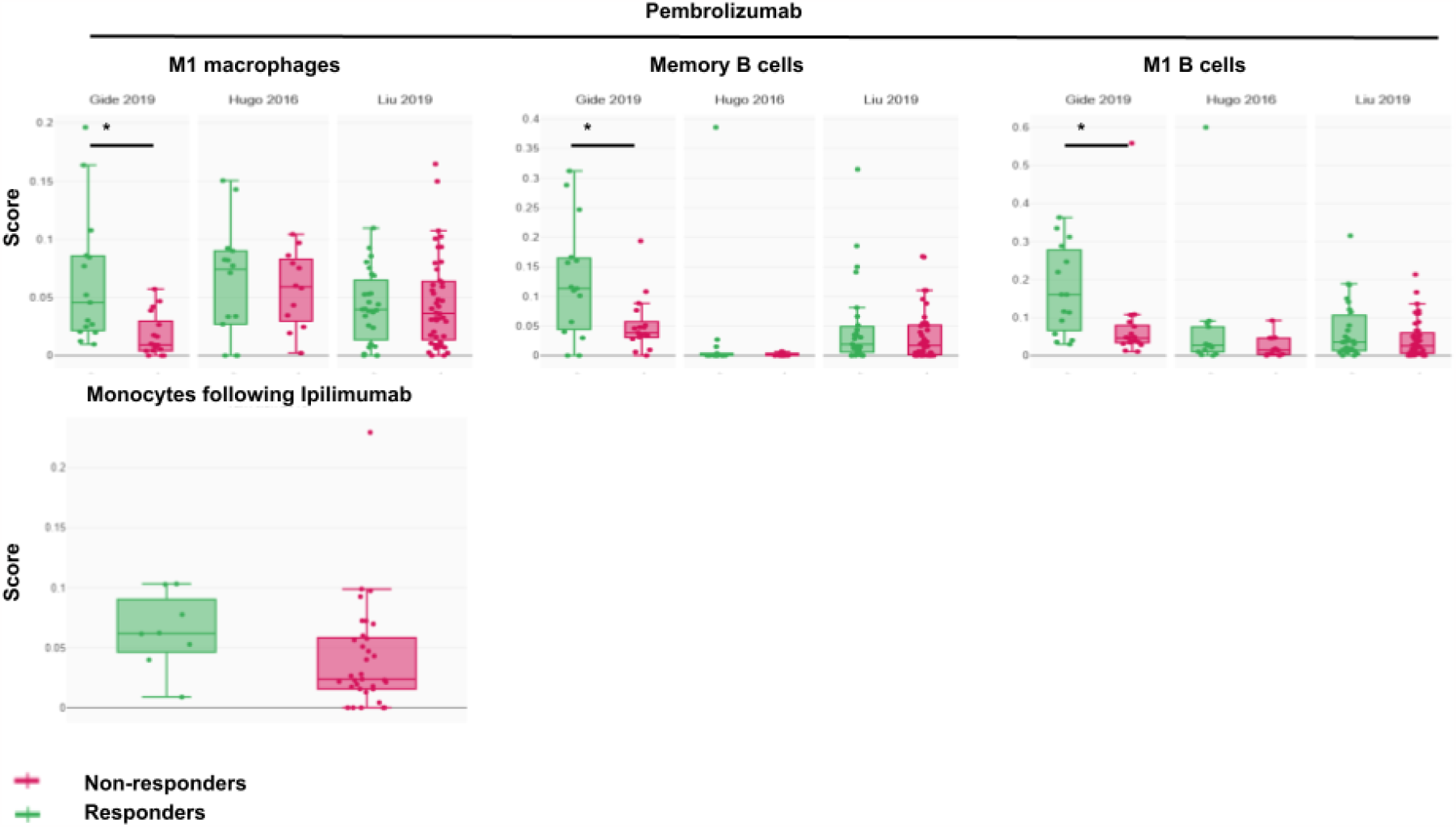
Immune infiltration in melanoma patients according to response to anti-PD1 or anti-CTLA4 monotherapies. 41 patients received Ipilimumab monotherapy. 160 patients received Nivolumab monotherapy.137 patients received Pembrolizumab monotherapy. Immune infiltration is measured using CRI iAtlas. p<0.05, Wilcoxon t-test.

**Supplementary figure 2).**
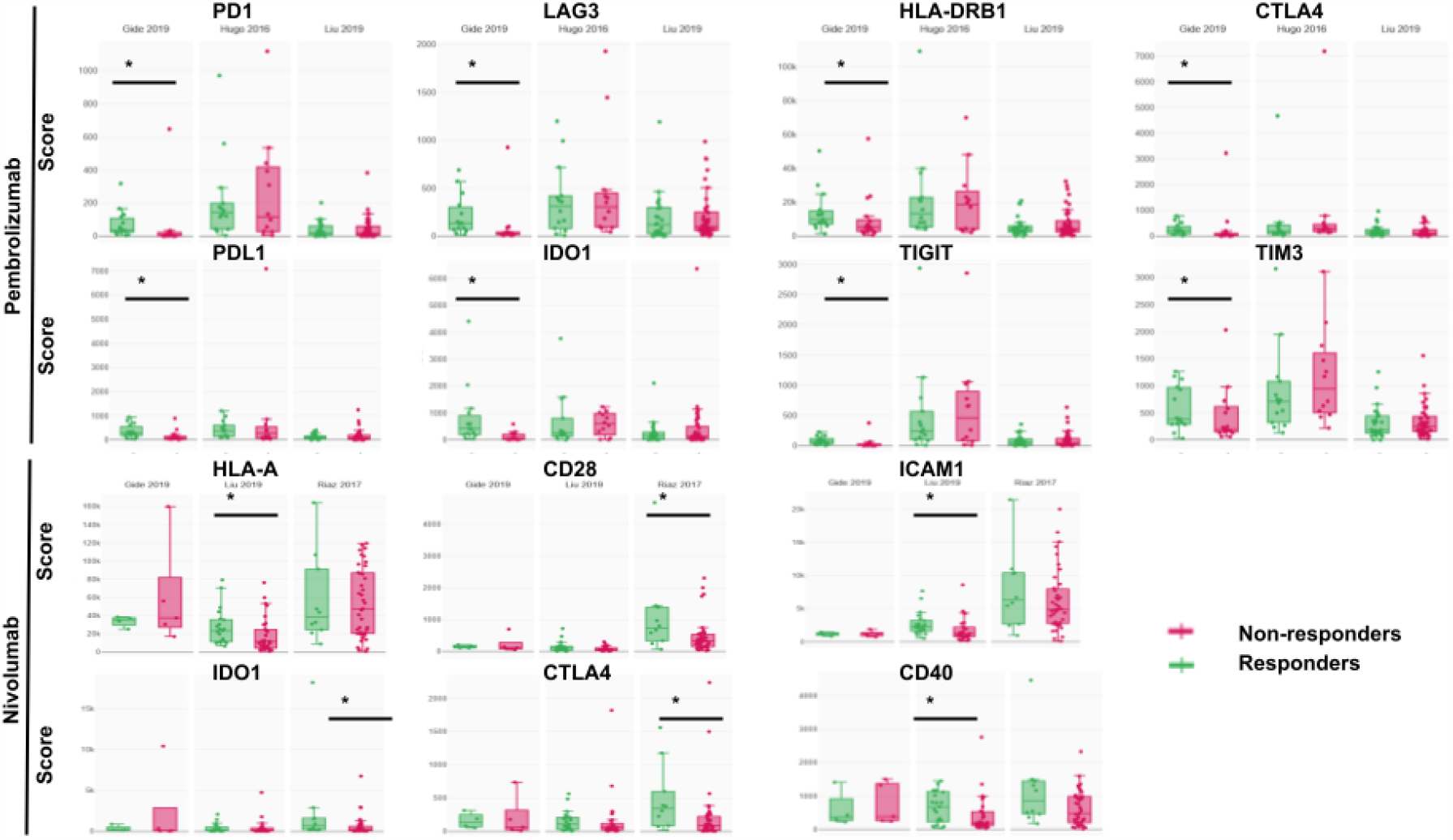
Immunomodulatory molecule expression in melanoma patients according to response to anti-PD1 or anti-CTLA4 monotherapies. 41 patients received Ipilimumab monotherapy. 160 patients received Nivolumab monotherapy.137 patients received Pembrolizumab monotherapy. Immunomodulatory molecule expression is measured using CRI iAtlas. p<0.05, Wilcoxon t-test.

